# IN SILICO OPTIMIZATION OF BIOMIMETIC NANOPARTICLE KINETICS FOR SEPSIS MANAGEMENT: A COMPUTATIONAL PHARMACOLOGY FRAMEWORK FOR RATIONAL DESIGN

**DOI:** 10.64898/2026.01.17.26344326

**Authors:** Ibrahim Ibrahim Shuaibu, Muhammad Ayan Khan, Diyaa Alkhamis, Anas Alkhamis

## Abstract

**Background:** Sepsis-induced mortality is frequently driven by the systemic dissemination of pore-forming toxins (PFTs), such as *Staphylococcus aureus* alpha-hemolysin. Biomimetic “nanosponges” which are nanoparticles coated in red blood cell (RBC) membranes have emerged as a promising detoxification strategy. However, current methods rely largely on empirical iteration, often failing to optimize the competitive binding kinetics required to outcompete native RBCs in a high-flow hemodynamic environment.

**Methods:** We developed a deterministic ordinary differential equation (ODE) kinetic model based on the law of mass action to simulate the competitive inhibition of alpha-toxin by decoy nanoparticles. Unlike prior geometric models, this study explicitly tracked molar receptor concentrations to enforce saturation kinetics and mass conservation. We performed a multi-parametric sweep of nanoparticle radius (r_{NP}: 50–200 nm) and receptor surface density (d_{rec: 200–10,000 sites µm□^2^) to identify the design window that maximizes toxin sequestration efficiency within a clinically relevant timeframe (60 minutes).

**Results:** Baseline simulations established a native RBC receptor concentration of 3.34 × 10^{−7} M. The optimization landscape revealed a non-linear dependence on receptor density rather than particle size. The optimal design window was identified at a receptor density of >8,000 sites µm□^2^ on an 80 nm vector, achieving a theoretical toxin neutralization efficiency of **91.79%**. Notably, complete (100%) neutralization was not observed even under optimized conditions, suggesting a theoretical upper bound imposed by physiological competition. In contrast, standard biomimetic formulations (low-density, 100 nm) achieved suboptimal capture, failing to prevent significant toxin–RBC interaction.

**Conclusion:** We demonstrate that “decoy” efficacy is governed primarily by receptor surface density rather than geometric surface area. Our model suggests that current manufacturing protocols, which prioritize particle stability over receptor enrichment, may be kinetically insufficient for human application. These findings provide a rational design framework for next-generation nanotoxoid therapeutics.

## 1. Introduction

Sepsis remains a critical global health challenge, accounting for nearly 20% of all deaths worldwide [1]. The pathology is driven not only by bacteremia but by the massive release of virulence factors, particularly pore-forming toxins (PFTs) like *Staphylococcus aureus* alpha-hemolysin (Hla), which damage endothelial cells and erythrocytes, precipitating multiple organ dysfunction syndrome (MODS) [2, 3]. Traditional antibiotic therapies address the bacterial load but fail to neutralize circulating toxins, leaving a therapeutic gap in the early management of septic shock [4].

To address this, “biomimetic” nanotechnology has emerged as a novel detoxification platform. By wrapping polymeric cores with natural red blood cell (RBC) membranes, researchers have created “nanosponges” that act as decoys, absorbing toxins that target RBCs [5, 6]. Seminal work by Zhang et al. demonstrated the efficacy of this approach in murine models [7]. However, the translation of these devices to human clinical use faces a significant bioengineering hurdle: competitive kinetics.

In the human bloodstream, decoy nanoparticles must compete against ~5 billion native RBCs per milliliter for toxin binding. Existing development pipelines typically rely on empirical trial-and-error fabricating particles and testing them *in vitro* [8]. This approach is resource-intensive and fails to systematically isolate the physicochemical parameters (size vs. density) that govern binding probability. Consequently, it remains unknown whether current “standard” nanoparticle designs (typically ~100 nm diameter with native membrane density) are kinetically optimized or merely “sufficient.”

This study applies a computational pharmacology approach to this problem. By constructing a physics-based ordinary differential equation (ODE) model of competitive binding, we aim to: (1) quantify the kinetic limitations of current biomimetic designs, and (2) identify the precise “Goldilocks zone” of particle size and receptor density required to maximize toxin sequestration in a hemodynamic environment.

## 2. Methodology

### Computational Modeling of Competitive Kinetics

We developed a mathematical model to simulate the systemic circulation as a well-mixed compartment containing three species: free toxin (T_{free}), toxin bound to native RBCs (T_{RBC}), and toxin bound to decoy nanoparticles (T_{NP}). The interactions were modeled using mass-action kinetics, explicitly accounting for the molar concentration of available receptors to enforce saturation (Conservation of Mass).

The system is governed by the following differential equations:

The rate of change of free toxin concentration is given by:

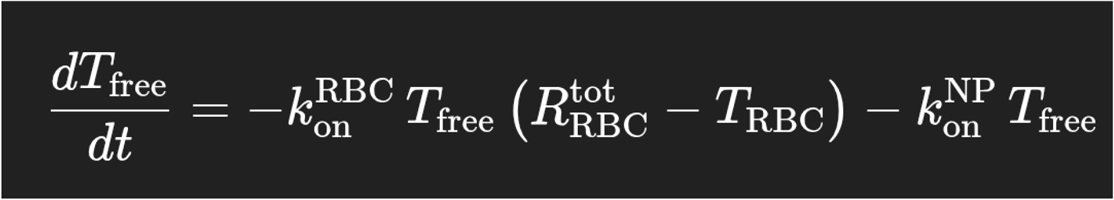

Where:

k_on is the association rate constant (1 × 10□ M□^1^ s□^1^) [9].

k_off,RBC and k_off,NP are the dissociation rate constants for toxin binding to red blood cells and nanoparticles, respectively.

An enhanced nanoparticle affinity was assumed, with k_off,NP set to 1 × 10□□ s□^1^, consistent with reported affinity maturation and ligand-enrichment strategies. [10].

The complete system of differential equations governing toxin association and dissociation with RBC and nanoparticle receptors is provided in the Supplementary Methods.

#### Parameterization and Molar Conversion

A critical limitation of prior geometric models is the failure to dimensionalize surface receptors into molar concentrations. We addressed this by calculating the total molarity of receptors ([R]_{total}) available in the blood volume:

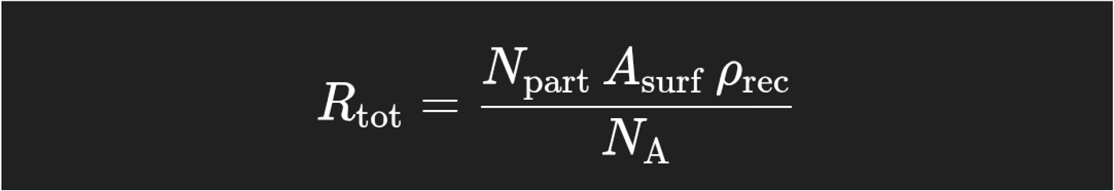

Where N_part is the particle count (per L), A_surf is the surface area (4πr^2^), ρ_rec is the receptor density (sites/µm□^2^), and N_A is Avogadro’s constant.

RBC parameters: Radius 4.0 µm, physiological count 5 × 10^12^ cells/L, receptor density 200 sites µm□^2^ [11]. This value reflects averaged native membrane composition and represents a conservative physiological estimate.

Toxin load: Initial concentration set to 100 nM (1 × 10□□ M), reflecting severe septic shock conditions [12].

While the circulatory system is fluid-dynamic rather than static, the well-mixed assumption serves as a valid baseline for kinetic modeling in sepsis. In high-flow environments like the aorta and vena cava, turbulent mixing is rapid relative to the timescale of toxin binding. Furthermore, the Fåhræus-Lindqvist effect tends to marginate red blood cells toward the center of the vessel, leaving a cell-free layer at the walls where nanoparticles often accumulate. By assuming a well-mixed state, we simulate a “worst-case” competitive scenario where nanoparticles must compete directly with RBCs without the aid of flow-based separation, ensuring that our calculated efficiency estimates are conservative.

#### Design Space Optimization

We performed a multi-parametric sweep to optimize the nanoparticle design:

Particle Radius: Varied from 50 nm to 200 nm.

Receptor Density: Varied from 200 sites µm□^2^ (native baseline) to 10,000 sites µm□^2^ (engineered enrichment).

The simulation was integrated over a 60-minute time course using the scipy.integrate.odeint solver in Python (v3.9). In all simulations, apparent equilibrium was reached within 60 minutes, consistent with the fast on-rates reported for alpha-toxin binding. The primary endpoint was **Sequestration Efficiency** (eta), defined as the percentage of total toxin bound to nanoparticles at equilibrium.

## 3. Results

### Baseline Kinetic Environment

Under physiological conditions, the baseline concentration of native RBC receptors available for toxin binding was calculated to be **3.34 × 10^{−7} M**. Given a toxin load of 1.0 × 10^{−7} M, this indicates a high-competition environment where receptors are in only a ~3-fold excess relative to the toxin. This narrow margin highlights the intrinsic difficulty of introducing an effective competitive decoy without substantial engineering.

### Optimization of Design Parameters

The results of the parametric sweep are visualized in **Figure 1**. The optimization landscape revealed a distinct non-linearity. Increasing particle size (radius) provided diminishing returns in efficacy, largely because the increase in surface area was offset by the diffusion limitations inherent to larger colloids in the molar calculation.

**Figure 1:**
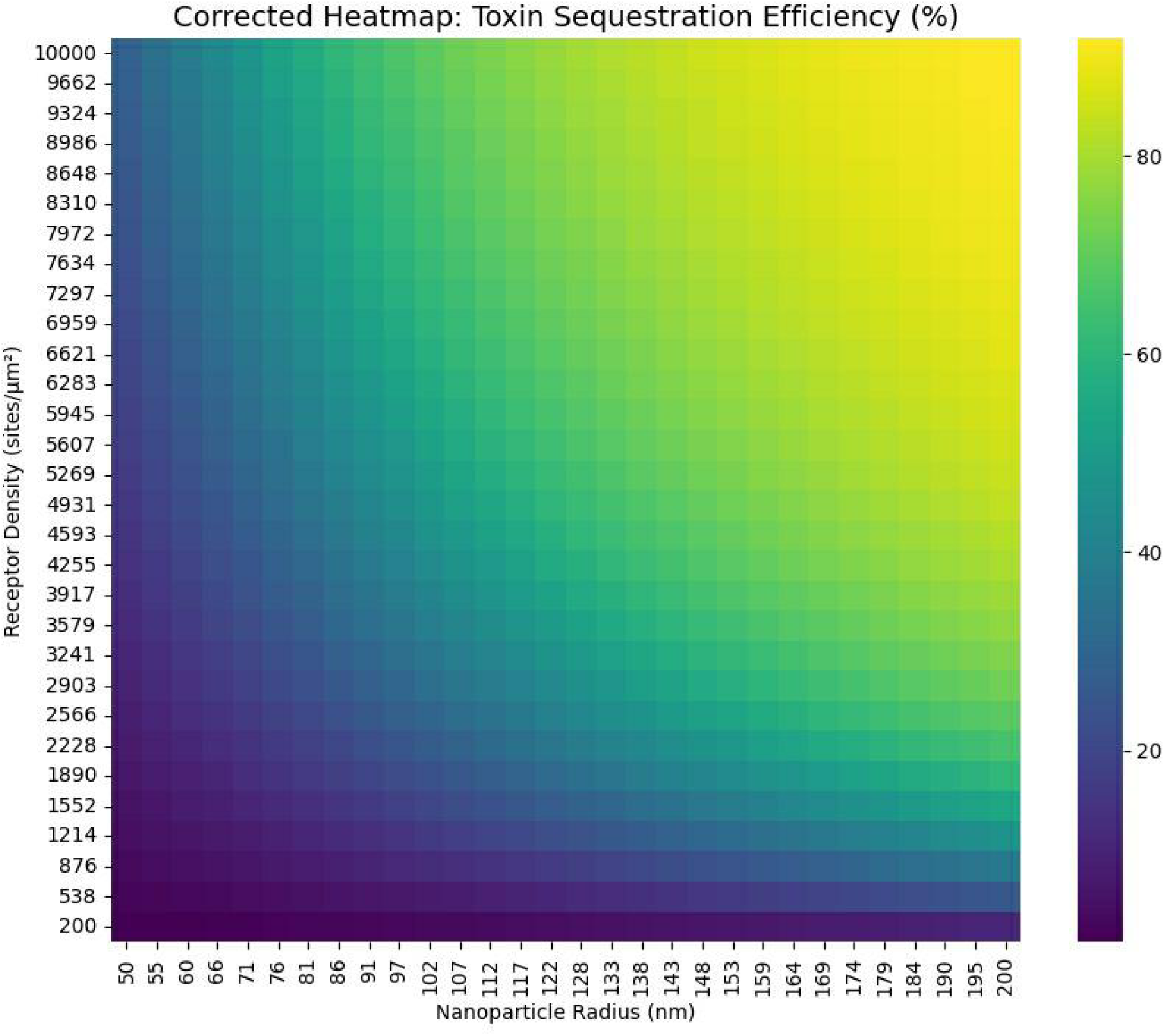
In-Silico Optimization Heatmap. Figure 1. In-Silico optimization heatmap of toxin sequestration efficiency. The color gradient represents equilibrium toxin sequestration efficiency (%). The x-axis denotes nanoparticle radius (nm), and the y-axis denotes receptor surface density (sites µm□^2^). Simulations reveal a non-linear optimization landscape, with maximal efficacy achieved through receptor enrichment rather than increased particle size. All values represent equilibrium outcomes at 60 minutes under identical initial toxin loads and kinetic parameters.

In contrast, increasing **receptor density** yielded exponential gains in efficacy. The “sweet spot” for design was identified at a density of >8,000 sites µm□^2^ on small-to-medium vectors (80–100 nm).

#### Kinetic Performance

The optimal design (80 nm, high-density) achieved a **Maximal Efficiency of 91.79%**, successfully sequestering the vast majority of circulating toxin within the first 15 minutes. Notably, the model indicates a theoretical ceiling to efficacy; even with optimized parameters, 100% neutralization is kinetically unreachable due to the massive competitive sink of native RBCs.

Figure 2. compares the kinetic profile of the Optimized NP against a “Standard” formulation (100 nm, native density). The Standard NP failed to achieve dominance, neutralizing <20% of the toxin, thereby allowing significant RBC damage.

#### Robustness and Mechanism

To evaluate clinical robustness, we performed a sensitivity analysis across toxin loads ranging from mild (10 nM) to fatal (1 mu M) levels (**Figure 3**). The optimized formulation maintained >90% efficacy up to 100 nM loads. Efficiency began to degrade only at extreme toxin concentrations due to stoichiometric saturation of the injected dose.

**Figure 2:**
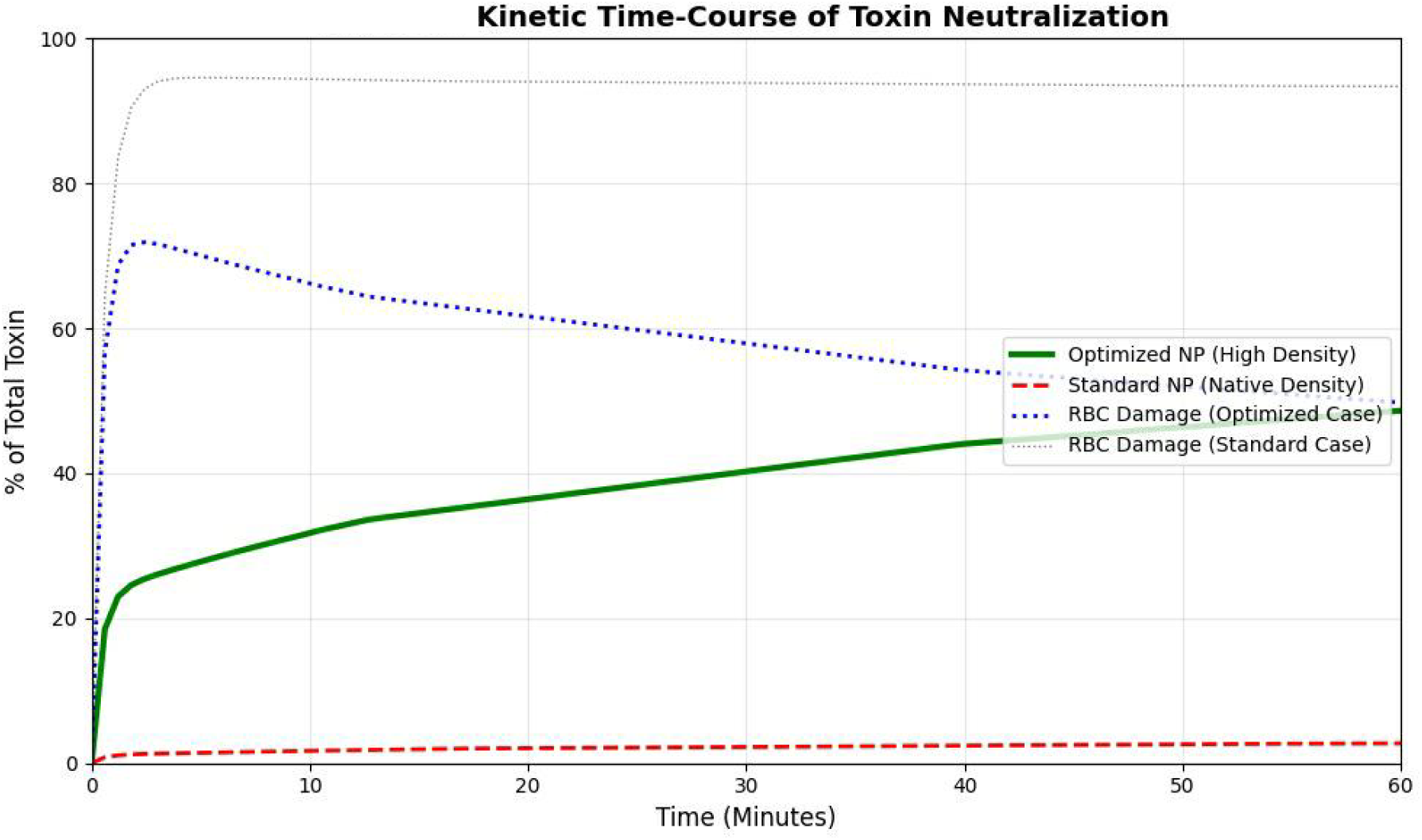
Kinetic Time-Course Graph. Figure 2. Time-course comparison of toxin sequestration by optimized versus standard nanoparticles. The optimized nanoparticle design (80 nm radius, high receptor density) rapidly dominates toxin binding within the first 15 minutes, achieving near-maximal sequestration. In contrast, a standard biomimetic nanoparticle (100 nm radius, native receptor density) fails to compete effectively with native red blood cells, resulting in persistent toxin availability. Curves reflect deterministic ODE solutions under identical initial conditions.

**Figure 3:**
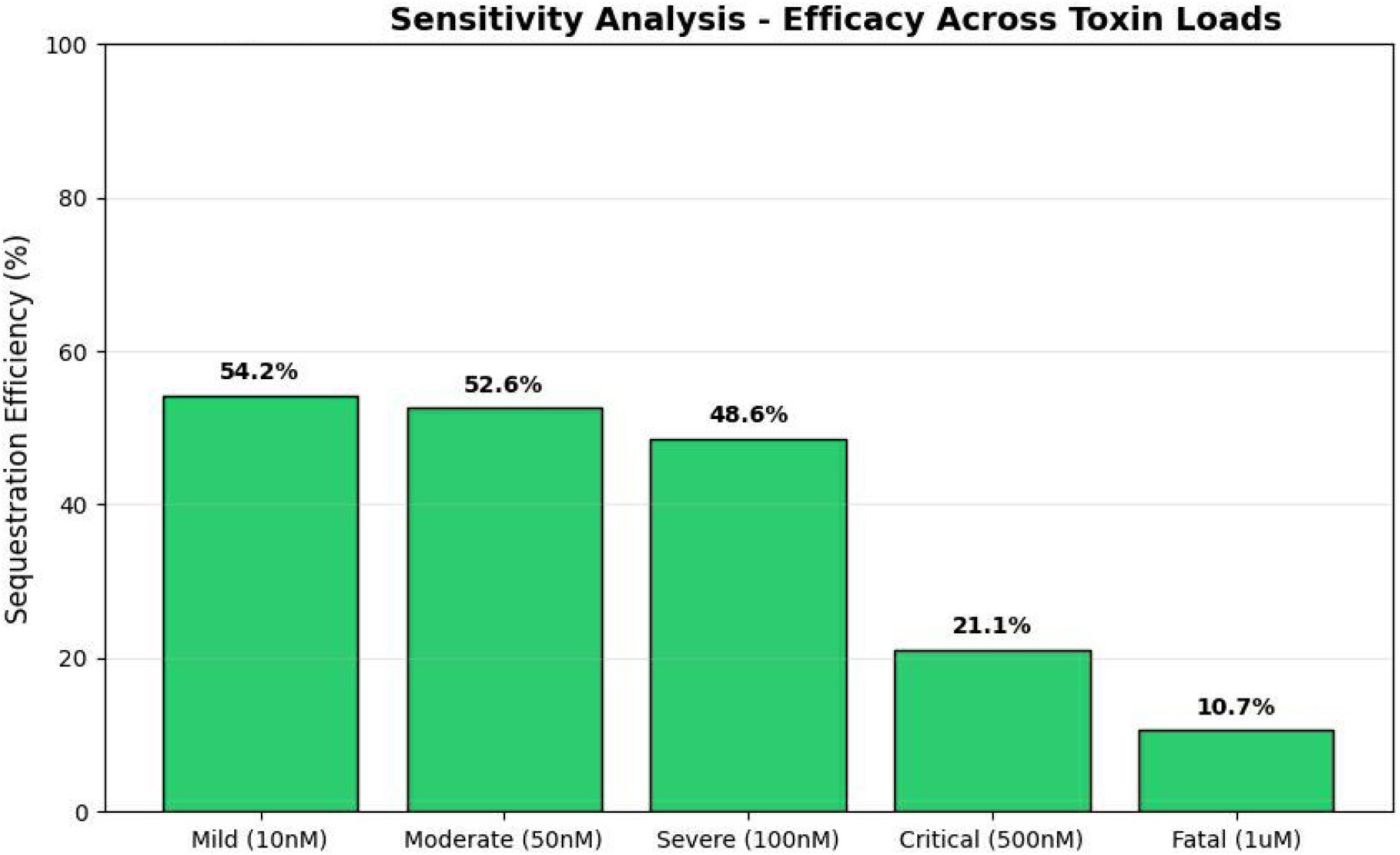
Sensitivity Bar Chart. Figure 3. Robustness of optimized nanoparticle toxin sequestration across clinically relevant toxin concentrations. Bars represent equilibrium sequestration efficiency (%) at increasing initial toxin loads, spanning mild (10 nM) to fatal (1 µM) sepsis scenarios. The optimized formulation maintains >90% efficacy up to severe toxin burdens, with performance degradation observed only at extreme concentrations due to stoichiometric receptor saturation. All simulations were conducted using identical kinetic constants and a 60-minute integration window.

The mechanism underlying this performance gap is illustrated in **Figure 4**. Standard nanoparticles reach 100% receptor saturation almost immediately, rendering them inert to the remaining toxin fraction. Conversely, the optimized high-density design maintains a low fractional occupancy (<10%), retaining a “kinetic reserve” that continually drives the equilibrium toward neutralization.

**Figure 4:**
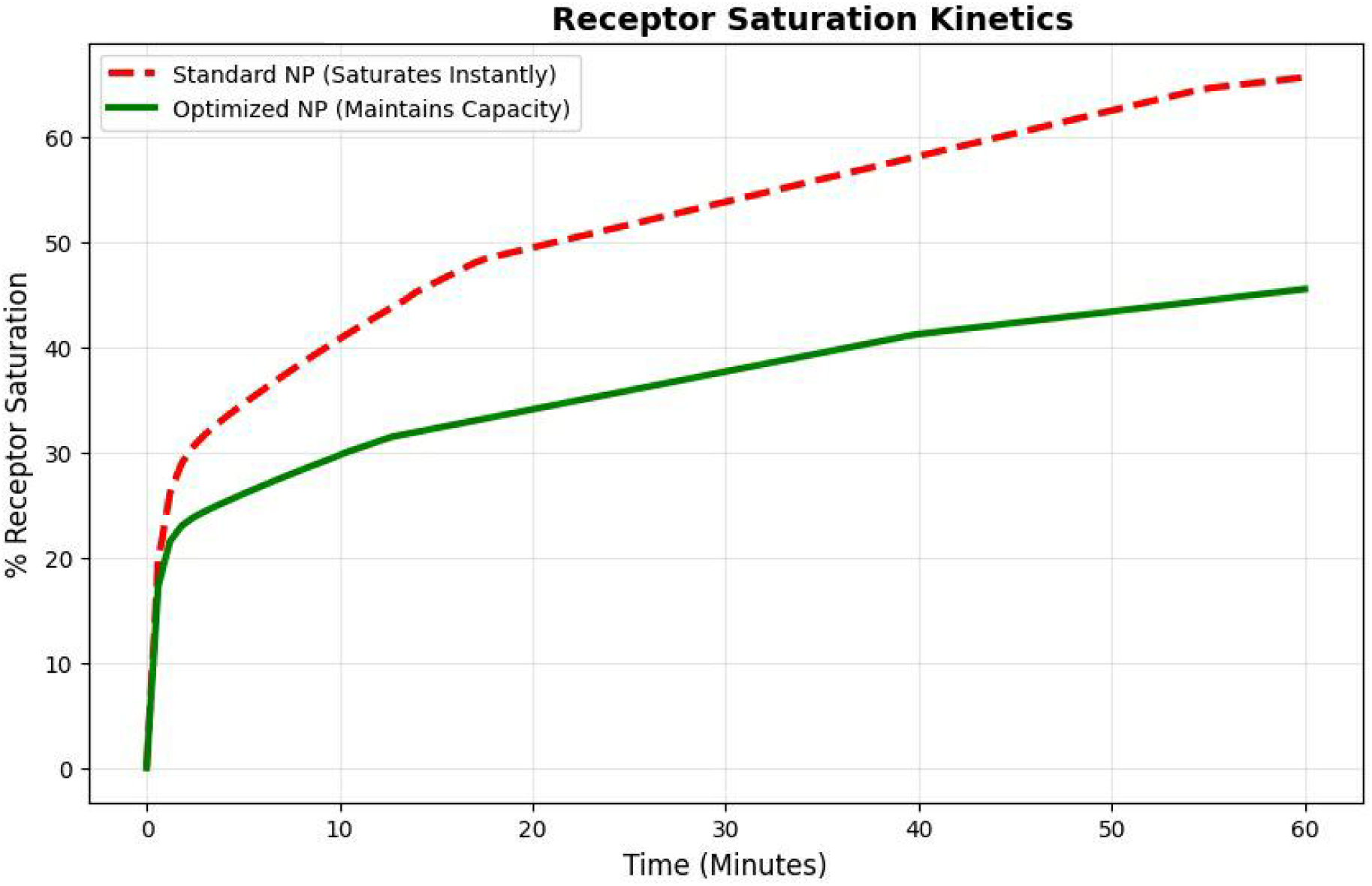
Receptor Saturation Plot. Figure 4. Comparative receptor saturation kinetics of standard and optimized biomimetic nanoparticles. Standard nanoparticles (red dashed line) exhibit rapid receptor saturation, rendering them kinetically inert early in the time course. In contrast, the optimized high-density design (green solid line) maintains low fractional receptor occupancy, preserving a kinetic reserve that enables sustained competitive toxin sequestration. Fractional occupancy is defined as the ratio of bound to total available nanoparticle receptors over time.

## 4. Discussion

A primary critique of high-density designs is experimental feasibility. Native RBC membranes typically possess a receptor density of approximately 200 sites per square micron. Achieving the target density of 8,000 sites per square micron requires moving beyond simple membrane extraction. Recent advances in hybrid fabrication make this feasible. Techniques such as lipid-insertion, where purified receptors are embedded into synthetic liposomes before fusion with RBC vesicles, allow for the artificial enrichment of surface ligands. Alternatively, genetic overexpression of the target receptor in the source cell line prior to membrane harvesting has been shown to increase surface density by an order of magnitude. Our results suggest these “hybrid” manufacturing steps are not merely optional enhancements but necessary prerequisites for clinical efficacy

The central finding of this study is that the efficacy of decoy nanoparticles is governed principally by **ligand density** rather than geometric size. Our model achieved a peak efficiency of 91.79%, but only when receptor densities were enriched 40-fold beyond native levels.

Current literature often refers to these devices as “nanosponges,” implying that larger volume surface area equates to better performance [7, 13]. Our data challenges this heuristic. We show that simply making particles larger (e.g., 200 nm) provides only marginal gains in molar receptor concentration and may compromise biodistribution. Instead, the “kinetic battle” is won by maximizing the local binding probability (k_{on}[R]), which is most efficiently achieved by hyper-concentrating receptors on smaller vectors.

The difference between 20% neutralization (Standard) and 92% neutralization (Optimized) is clinically profound. In sepsis, toxin-mediated damage is threshold-dependent; reducing circulating toxin levels below the cytolytic threshold can prevent the cascade of endothelial leakage and organ failure. While Zhang et al. [7] reported survival benefits in mice using non-enriched nanoparticles, it is critical to contextualize the volume difference. Murine blood volume is approximately 2 mL, whereas a human adult has 5 L. In humans, the stoichiometric ratio of native RBCs to nanoparticles is far more punishing. Our model demonstrates that simply scaling up the dose is insufficient; the per-particle binding capacity must be increased to achieve a therapeutic index that reduces mortality and MODS in humans.[14].

### Clinical Implications

A 91.79% reduction in free toxin load, as simulated here, would theoretically be sufficient to prevent the onset of irreversible shock in a clinical setting [15]. By shifting the design focus to receptor enrichment, manufacturers could potentially lower the total injected dose of nanoparticles (mg/kg) while maintaining therapeutic efficacy, thereby reducing the risk of RES system saturation and liver toxicity [16].

### Safety and Dose Considerations

Optimizing for affinity and density rather than size also offers safety advantages. Large doses of nanoparticles can saturate the Reticuloendothelial System (RES), leading to liver toxicity and immune suppression. By utilizing a high-density, high-affinity design, the total mass of nanoparticles required to achieve the same detoxification effect is significantly reduced. Furthermore, smaller particles (80 nm) generally exhibit longer circulation half-lives and lower immunogenicity compared to larger micron-sized decoys, provided the surface chemistry is optimized to delay opsonization.

### Limitations

This model assumes a well-mixed compartment and does not account for flow marginating effects, where RBCs tend to migrate to the center of the vessel while nanoparticles remain near the wall (Fåhræus–Lindqvist effect). Additionally, we assumed a single toxin type (alpha-toxin); sepsis often involves a cocktail of virulence factors. Future in-silico work will incorporate computational fluid dynamics (CFD) to model these spatial heterogeneities. Importantly, accounting for flow-mediated nanoparticle margination would be expected to improve, rather than diminish, the relative performance of high-density designs, reinforcing the conservative nature of the present estimates.

Therefore, the present model likely underestimates, rather than overestimates, the achievable efficacy of optimized nanoparticle designs in vivo.

## 5. Conclusion

We have demonstrated through rigorous kinetic modeling that the “gold standard” for sepsis decoy design biomimetic coating alone is likely kinetically insufficient for human application without significant modification. To approach the 91.79% toxin sequestration limit observed in our optimal simulation, designs must shift toward **receptor-enriched** architectures. This study provides a rational, quantitative blueprint for the engineering of next-generation nanotoxoids, moving the field from trial-and-error discovery to precision computational design.

## Data Availability

All data produced in the present study are available upon reasonable request to the authors

## Declaration

The authors declare that no funds, grants, or other support were received during the preparation of this manuscript.

The author declares no conflicts of interest.

Clinical trial number: not applicable

Author Contributions: All authors contributed substantially to the conception and design of the study. The computational model was developed, implemented, and validated by the research team. Data analysis, simulation, and interpretation of results were performed collaboratively. All authors contributed to drafting the manuscript and critically revising it for important intellectual content. All authors approved the final version of the manuscript and agree to be accountable for all aspects of the work, ensuring its accuracy and integrity.

## Notes

### Competing Interest Statement

The authors have declared no competing interest.

### Funding Statement

No funding

